# Diagnostic performance and kinetics of hepatitis E viral RNA and IgM antibody test positivity in a genotype 1 outbreak in South Sudan

**DOI:** 10.1101/2025.04.03.25325193

**Authors:** Aybüke Koyuncu, Robin Nesbitt, Catia Alvarez, Kinya Vincent Asilaza, Joseph Wamala, Melat Haile, Etienne Gignoux, Manuel Albela, Emily S. Gurley, Frederick Beden Loro, Duol Biem, Monica Rull, John Rumunu, Iza Ciglenecki, Isabella Eckerle, Andrew S. Azman

## Abstract

**Background:** Diagnostics are essential for understanding hepatitis E epidemiology, but the field performance of available tests remains unclear. We evaluated the performance of PCR, IgM ELISA, and the Assure HEV IgM rapid diagnostic test (RDT) during a HEV genotype 1 outbreak and assessed the duration of viremia and antibodies responses.

**Methods:** We used data from enhanced surveillance at a health facility in Bentiu internally displaced persons camp, South Sudan (March-December 2022). Suspected hepatitis E cases underwent testing with all three diagnostics at enrolment, with a follow-up sample collected as part of a vaccine effectivness study. We used a latent class model to estimate test performance and accelerated failure time models to estimate time from jaundice onset to a negative test for PCR and ELISA.

**Findings:** Among 893 suspected cases, test sensitivity declined with time from jaundice onset. Within 30 days of jaundice onset, PCR sensitivity was 73% (95% Credible Interval (CrI) 27, 90), compared to 86% for RDT (95% CrI: 74, 93), and 95% for ELISA (95% CrI: 91, 98). Specificity was high across tests: PCR at 98% (95% CrI: 98, 99), RDT at 95% (95% CrI: 93, 96), and ELISA at 95% (95% CrI: 93, 96). Median time from jaundice onset to negative test was 19 days (95% CI: 17, 21) for PCR and 113 days (95% CI: 87, 163) for ELISA.

**Interpretation:** The Assure RDT showed higher sensitivity than PCR and similar specificity to ELISA, supporting its use in surveillance. Care seeking delays can greatly influence the interpretation of diagnostic tests.

## Background

Hepatitis E causes an estimated 3 million symptomatic cases of acute viral hepatitis each year with case fatality risks of up to 25% often reported among pregnant women (1,2), though the true burden is unknown. Large outbreaks, with thousands of cases, caused by hepatitis E virus (HEV) genotypes 1 and 2 occur due to fecal contamination of drinking water (3).

Understanding the true burden of hepatitis E is challenging due to lack of routine testing for hepatitis E among acute jaundice cases, particularly in low- and middle-income settings. A safe and effective vaccine for hepatitis E exists (Hecolin; Innovax, Xiamen, China) (4), but the World Health Organization (WHO) has identified gaps in burden of disease data as a barrier to widespread introduction of the vaccine (5). The vaccine is currently only recommended by WHO for use in outbreak settings (5).

Many settings where hepatitis E outbreaks occur lack resources and laboratory infrastructure to conduct confirmatory testing of suspected hepatitis E cases. Once a hepatitis E outbreak is suspected, biological samples from suspected cases are shipped to reference laboratories, often in other countries, for confirmation based on detecting HEV RNA using reverse transcriptase polymerase chain reactions (PCR) or IgM antibodies specific to HEV using ELISA. In a displaced persons camp in Niger, for example, health authorities were notified about an increase in acute jaundice syndrome cases in January 2017 but “in the absence of appropriate diagnostic capacities” serum samples were sent to Senegal for biological confirmation (6). Following confirmation of HEV, the outbreak was officially declared in April (6). The need to wait for confirmation from external laboratories slows down mobilization of resources for public health interventions that can interrupt transmission. These delays also decrease the potential impact of reactive vaccination campaigns, which have the greatest benefits in preventing cases and deaths when implemented in the early stages of an outbreak (7).

Rapid diagnostic tests (RDTs) for hepatitis E, which can detect IgM antibodies or viral antigen (2), require less laboratory infrastructure and can be conducted at point-of-care, but their field performance is not well described. In 2022, the WHO Strategic Advisory Group of Experts on In Vitro Diagnostics (SAGE IVD) conditionally added RDTs for HEV to WHO’s list of essential in vitro diagnostics, noting that although additional information on test performance is needed, RDTs can facilitate access to hepatitis E diagnostics in settings where PCR and ELISA are not accessible (8). Few validation studies have been performed for HEV RDTs and among those that have, they have generally been conducted in controlled laboratory settings with ideal positive and negative samples (e.g. samples already known to be positive or negative) and with HEV genotype 3 (9–11). To our knowledge, no performance data from real world conditions have been published, where antibody or antigen concentrations may vary and where collection and transport conditions can influence test results. Evidence on the field performance of RDTs can help accelerate their broader use in surveillance and outbreak response.

The natural history of HEV, viral and antibody kinetics and analytical test performance shape the real world performance of HEV diagnostic tests and can challenge our ability to measure their true sensitivity and specificity. Symptoms develop on average 30 days after infection (95% CI 24–36) lasting 1 to 6 weeks (2,12). Limited existing evidence suggests that HEV RNA concentrations peak shortly before symptom onset in blood and decay quickly (13), potentially before a case seeks care. After infection, IgM antibodies take time to develop though it is not clear how quickly and how often antibody-based tests may miss early infections (14). Furthermore, IgM antibodies can last for more than six months (15), which can lead to false-positive test results. Assessing the performance of new diagnostic tests without taking into account the imperfections of reference assays and patient characteristics could lead to severe biases in estimates of test performance.

Here, we leverage data from acute jaundice surveillance nested within a vaccine effectivness study during an outbreak of hepatitis E genotype 1 in South Sudan to generate estimates on the sensitivity and specificity of PCR, IgM ELISA, and IgM RDTs in a real world setting and shed new light on the time course of HEV viremia and antibodies after onset of jaundice.

## Methods

### Study setting

Bentiu Internally Displaced Persons camp was established in December 2013 and cases of hepatitis E started to be detected there in 2014 (16). In March 2022 the South Sudan Ministry of Health in partnership with Médecins Sans Frontières (MSF) implemented the first-ever reactive vaccination campaign with Hecolin (17), coupled with a vaccine effectivness study. As part of the study, MSF reinforced the use of clinical case definitions and conducted comprehensive testing for all suspected cases (16). Acute jaundice syndrome (AJS) was defined as acute (recent, new, or sudden) onset of yellow coloration of the whites of the eyes or skin, dark urine or pale clay stools. All cases of AJS were considered suspected cases of hepatitis E.

All suspected hepatitis E cases seeking care at the MSF hospital were identified by clinicians and referred to the study team after consultation or admission. Study staff explained the study objectives and, among suspected cases willing to participate, obtained consent for participation. Adults provided written informed consent while individuals under 18 years of age provided assent and their guardian provided written informed consent. Study staff asked all consenting suspected cases questions about symptoms, date of symptom onset, vaccination status, and sociodemographic variables. To characterize viral and antibody kinetics over time, study participants had a followup visit at least 2 weeks after their enrollment where a blood sample was collected.

At each visit laboratory technician collected a venous blood sample and prepared all specimens for testing, storage, and transport. Plasma was separated by centrifugation and aliquoted at the MSF hospital, then frozen at -20°C within 6 hours. All samples were transported from Bentiu first to MSF, Juba, South Sudan, then to the reference laboratory in Switzerland on flights with temperature loggers in cold chain using dry ice. Samples were were stored in Juba and Geneva in -80°C freezers.

### Laboratory methods

A trained laboratory technician conducted HEV IgM RDTs and liver function tests at the Bentiu hospital laboratory. The HEV IgM RDT (Assure, Genelabs Diagnostics, Singapore, Republic of Singapore) was performed on ∼35uL of venous blood. The kit was stored at ambient temperature and was performed according to manufacturer’s instructions by a trained laboratory technician in Bentiu. AST and ALT concentrations were measured in venous blood using an automated system (Reflotron) or SimplexTAS machine in the hospital laboratory.

PCR and IgM ELISA testing was conducted in a reference laboratory at the University Hospitals of Geneva. We used WANTAI HEV-IgM ELISA and WANTAI HEV-IgG ELISA (WE-7196 and WE-7296, Beijing Wantai Biological Pharmacy Ent.) to detect HEV antibodies in venous plasma following the package insert.

RNA was extracted from plasma samples using the NucliSens easyMAG instrument (BioMérieux), following the manufacturer’s instructions. We used real-time quantitative polymerase chain reaction (RT-qPCR) to detect HEV RNA (Mikrogen Diagnostik, Bavaria, Germany) with primers targeting both the HEV ORF2 and ORF3 (18).

### Statistical analysis

We first estimated the sensitivity of IgM RDT and IgM ELISA using PCR as the reference standard. We then developed a latent class model to use results from all three tests to simultaneously estimate the sensitivity and specificity of each test (19). We included each individual’s PCR, RDT, and IgM ELISA test results and modelled the combination of the three tests as a multinomial likelihood:

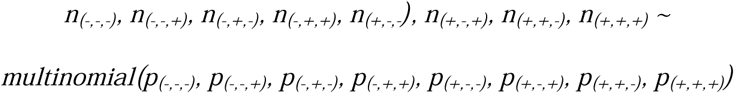

The probabilities for each sequence of test results were defined as follows:

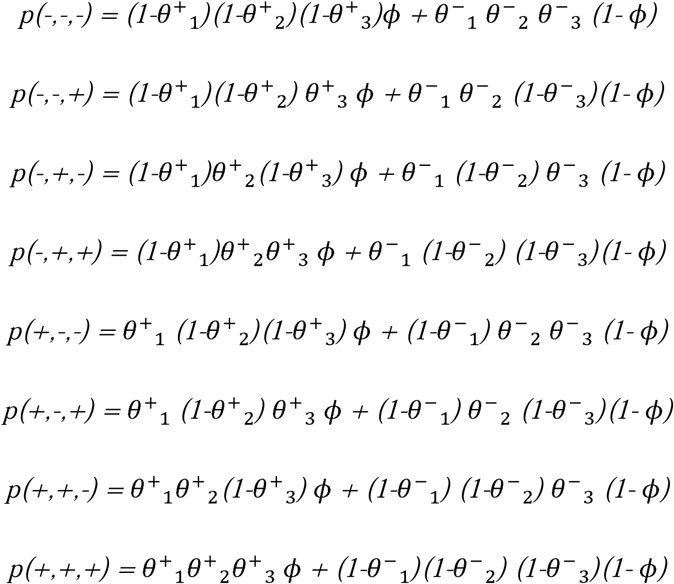

Where *ϕ* denotes the true probability of hepatitis E infection, *θ*^+^ denotes test sensitivity, *θ*^−^ denotes test specificity, and subscripts 1, 2, and 3 denote PCR, IgM RDT, and IgM ELISA. We included both test results at enrollment and at the follow-up visit in the model and marginalized out the unobserved IgM RDT result at follow-up. Priors and sensitivity analyses using alternate priors are described in Supplemental Tables 5 and 6.

We conducted inference in a Bayesian framework using Hamiltonian Markov Chain Monte Carlo as implemented in the STAN language. In the *unadjusted model*, we treated the sensitivity and specificity of each test as fixed across all individuals. We explored adjusted models where the logit sensitivity of each test varied by days between self-reported jaundice onset and clinic visit (linear or cubic spline with three degrees of freedom), age, the interaction between age and days between jaundice onset and clinic visit, and sex (Supplemental Table 3). We explored models with a constant risk of hepatitis E and those that allowed the risk to vary by month of AJS onset.

For each model, we ran 4 chains of 1,000 iterations after warm-up and assessed convergence among chains visually and using the Gelman-Rubin R-hat statistic (20). We used approximate leave-one-out cross-validation using Pareto smoothed importance sampling (21) to compare the predictive accuracy of each model and select the final model.

We used accelerated failure time models to obtain parametric and non-parametric estimates – accounting for the censored nature of the data – of the median time from jaundice onset to first negative test result among individuals who tested positive at enrollment, for PCR and IgM ELISA seperatley (22). We explored Weibull, log-normal, and gamma distributions for the survival curve and selected the distribution that minimized the Akaike Information Criterion (AIC). We quantified uncertainty using the 2·5^th^ and 97·5^th^ percentiles of 1,000 bootstrap draws. In sensitivity analyses we explored whether the time to first negative test varied by age (linear and categorical) and/or sex.

We conducted posterior retrodictive checks for both the final adjusted latent class model and AFT models to assess how well the final selected models could reproduce observed diagnostic test results. Code for all analyses and a minimal dataset are available at https://github.com/HopkinsIDD/hev-diagnostics-bentiu.

### Ethics

Ethical approval for the parent study was obtained from Médecins Sans Frontière (ERB #2167), the South Sudan Ministry of Health Research Ethics Board (RERB-MOH # 54/27/09/2022) and The Johns Hopkins Bloomberg School of Public Health Institutional Review Board (IRB00025966).

## Results

We identified 893 suspected hepatitis E cases between March and December 2022 with complete test results at enrollment (Table 1). Of these, 67% (N=602) had jaundice onset within 1 week and 90% (N=805) within 30 days of their clinic visit. Among suspected hepatitis E cases, 180 (20%) had viral RNA detected by PCR, 245 (27%) had IgM antibodies detected by RDT, and 236 (26%) had IgM antibodies detected by ELISA. Half of suspected cases were female (46%) and half were under 16 years old (49%). A higher proportion of suspected cases who tested positive on all tests were under 16 years old compared to suspected cases who had no viral RNA or IgM antibodies detected (67% versus 42%). 24 suspected cases were pregnant, of whom 23 were pan-negative and 1 was RDT-positive only. A majority of suspected cases had IgG antibodies detected by ELISA at the time of enrollment (85%). A higher proportion of pan-positive suspected cases returned for a follow-up visit with a blood draw compared to mixed and pan-negative suspected cases (46% versus 40% versus 39%). We excluded 1 suspected case missing jaundice onset date (0·11%) from subsequent analyses.

**Table 1.** Characteristics of suspected hepatitis E cases who presented to care at a health facility between March and December 2022 with complete testing data. Pan-positive = PCR, IgM ELISA, and IgM RDT+; Pan-negative = PCR, IgM ELISA and IgM RDT-

| Characteristic | Case type at enrollment |  |  |  |
| --- | --- | --- | --- | --- |
|  | Overall | Pan-positive | Mixed | Pan-negative |
| N (col %) | N=893 | N=169 | N=107 | N=617 |
| Female | 406 (45.5) | 77 (45.6) | 48 (44.9) | 281 (45.5) |
| Age, median (IQR) | 15.5<br>(5.9, 26.2) | 9.6<br>(4.9, 17.7) | 13.3<br>(5.4, 21.5) | 18.3<br>(6.9, 28.5) |
| Age |  |  |  |  |
| 0-5 | 192 (21.5) | 44 (26.0) | 26 (24.3) | 122 (19.8) |
| 6-15 | 241 (27.0) | 69 (40.8) | 34 (31.8) | 138 (22.4) |
| 16-39 | 379 (42.4) | 50 (29.6) | 38 (35.5) | 291 (47.2) |
| 40+ | 81 (9.1) | 6 (3.6) | 9 (8.4) | 66 (10.7) |
| Days since jaundice onset*, median (IQR) | 5.0<br>(3.0, 10.0) | 4.0<br>(3.0, 7.0) | 5.0<br>(3.0, 15.0) | 5.0<br>(3.0, 12.0) |
| Days since jaundice onset* |  |  |  |  |
| ≤1 week | 602 (67.4) | 132 (78.1) | 64 (59.8) | 406 (65.9) |
| >1 week to 2 weeks | 129 (14.4) | 30 (17.8) | 15 (14.0) | 84 (13.6) |
| >2 weeks to 1 month | 74 (8.3) | 6 (3.6) | 10 (9.3) | 58 (9.4) |
| >1 month to 2 months | 49 (5.5) | 1 (0.6) | 9 (8.4) | 39 (6.3) |
| >2 months | 38 (4.3) | 0 (0) | 9 (8.4) | 29 (4.7) |
| Anti-HEV IgG positive | 763 (85.4) | 169 (100) | 97 (90.7) | 497 (80.6) |
| Elevated ALT** | 135 (20.5) | 98 (72.1) | 4 (5.2) | 33 (7.4) |
| Elevated AST*** | 55 (17.5) | 8 (25.0) | 4 (10.3) | 43 (17.6) |
| Hospitalized**** | 57 (6.4) | 6 (3.6) | 13 (12.1) | 38 (6.2) |
| Died | 9 (1·0) | 0 (0) | 2 (1·9) | 7 (1·1) |
| Attended follow-up visit | 363 (40·6) | 77 (45·6) | 43 (40·2) | 243 (39·4) |

\*Missing for 1 suspected case
\*\*Missing for 233 suspected cases. For males, ALT ≥ 41 units per liter was considered elevated. For females, ALT >32 units per liter was considered elevated (23,24).
\*\*\*Missing for 578 suspected cases. AST above 40 units per liter was considered elevated.
\*\*\*\*Missing for 1 suspected case

### Diagnostic performance

Using PCR as the reference standard, the sensitivity of IgM RDT was 94·4% (95% Confidence interval (CI): 90·0, 97·3) and specificity was 89·5% (95% CI: 87·0, 91·6) (Table 2). The sensitivity and specificity of IgM RDT compared to IgM ELISA was 88·1% (95% Confidence interval (CI): 83·3, 92·0) and 94·4% (95% CI: 92·3, 96·0), respectively.

**Table 2.** Sensitivity and specificity of rapid diagnostic tests (RDT) compared to reverse transcription polymerase chain reaction (PCR) and enzyme-linked immunosorbent assays (ELISA).

| Gold standard |  |  | Metric (95% CI) |  |
| --- | --- | --- | --- | --- |
| Test results | PCR+ | PCR- | Sensitivity | Specificity |
| **IgM RDT+** | 170 | 75 | 94·4% | 89·5% |
| **IgM RDT-** | 10 | 637 | (90·0, 97·3) | (87·0, 91·6) |
|  | IgM ELISA+ | IgM ELISA- | Sensitivity | Specificity |
| **IgM RDT+** | 208 | 37 | 88·1% | 94·4% |
| **IgM RDT-** | 28 | 619 | (83·3, 92·0) | (92·3, 96·0) |

When taking into account all three HEV-specific tests, we estimated that 24.8% (95% CI: 22.2, 27.6) of suspected cases were true HEV infections. Using a latent class model, we estimate the average specificity of PCR was 98.2% (95% CrI: 97.6, 98.7), IgM RDT was 94.7% (95% CrI: 93.4, 95.9), and IgM ELISA to be 94.6% (95% CrI: 93.0, 95.9) (Table 3). Models allowing for sensitivity to decrease with increasing care-seeking delays were well supported by the data (Supplemental Table 3). When averaged across observed careseeking delays up to 30 days, PCR had the lowest sensitivity (73.4%, 95% CrI 27.2, 90.0), followed by IgM RDT (86.2%, 95% CrI: 74.0, 92.5) and IgM ELISA (95.2%, 95% CrI: 90.9, 97.7) (Table 3). The median sensitivity of PCR was 89.2% among individuals presenting for care the day of jaundice onset (95% CrI: 84.2, 92.7), 60.1% after two weeks (95% CrI 51.0, 68.8) and 20.2% after one month (95% CrI: 10.4, 34.7) (Figure 1). The sensitivity of RDT was 90.2% among those presenting for care the day of jaundice onset (95% CrI: 85.9, 93.8), 83.4% after two weeks (95% CrI: 77.3, 88.8) and 72.7% after one month (95% CrI: 62.8, 81.3). The sensitivity of IgM ELISA was 96.5% among those presenting for care the same day as jaundice onset (95% CrI: 94.1, 97.9), 94.5% after two weeks (95% CrI: 91.3, 96.7) and 91.4% after one month (95% CrI: 86.3, 94.7). In models that allowed the decline in sensitivity by careseeking delays to vary by age group, the sensitivity of all diagnostic tests, most notably those based on IgM, declined more quickly for individuals under 5 years of age compared to over 5 years (Supplemental Figure 6). Alternative models, including those with different defintions for symptom onset and different priors led to similar estimates of test performance (Supplemental Table 4 and 6).

**Figure 1.**
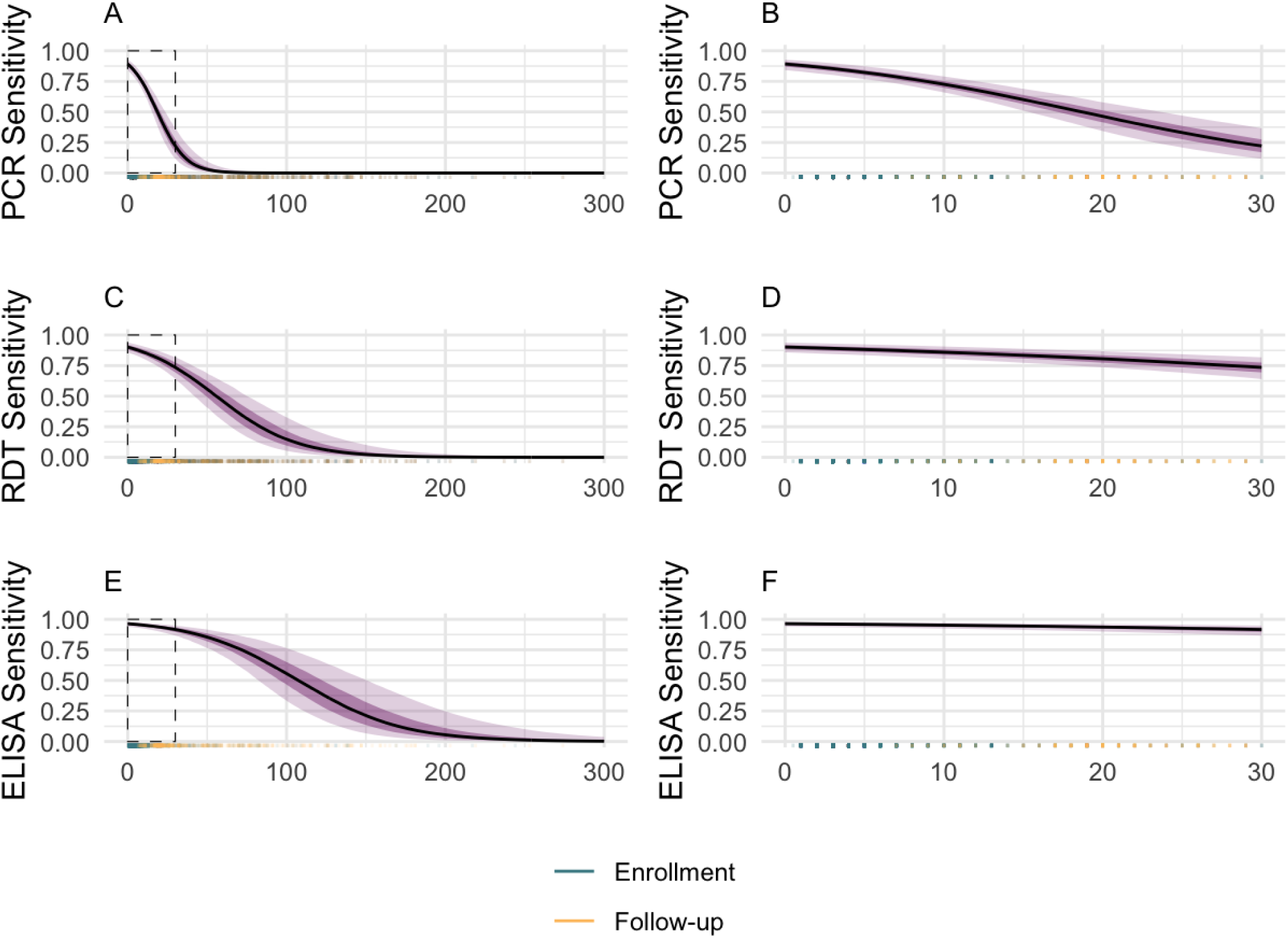
Sensitivity and specificity of diagnostic tests for detecting hepatitis E infections as a linear function of days between jaundice onset and clinic visit. A: PCR sensitivity across all careseeking delays with box around the first 30 days between jaundice onset and clinic visit; B: PCR sensitivity in the first 30 days between jaundice onset and clinic visit; C: IgM RDT sensitivity across all careseeking delays with box around the first 30 days between jaundice onset and clinic visit; D: IgM RDT in the first 30 days between jaundice onset and clinic visit; E: IgM ELISA sensitivity across all careseeking delays with box around the first 30 days between jaundice onset and clinic visit; F: IgM ELISA in the first 30 days between jaundice onset and clinic visit; Rug plot showing frequency of observed values. Line indicates median, dark purple indicates 20^th^ and 80^th^ quantiles, and light purple indicates 2·5^th^ and 97·5^th^ quantiles. C: Average sensitivity and specificity by test with 95% credible interval. Logit sensitivity varied linearly by days between self-reported jaundice onset and clinic visit, separately for each test.

**Table 3.**
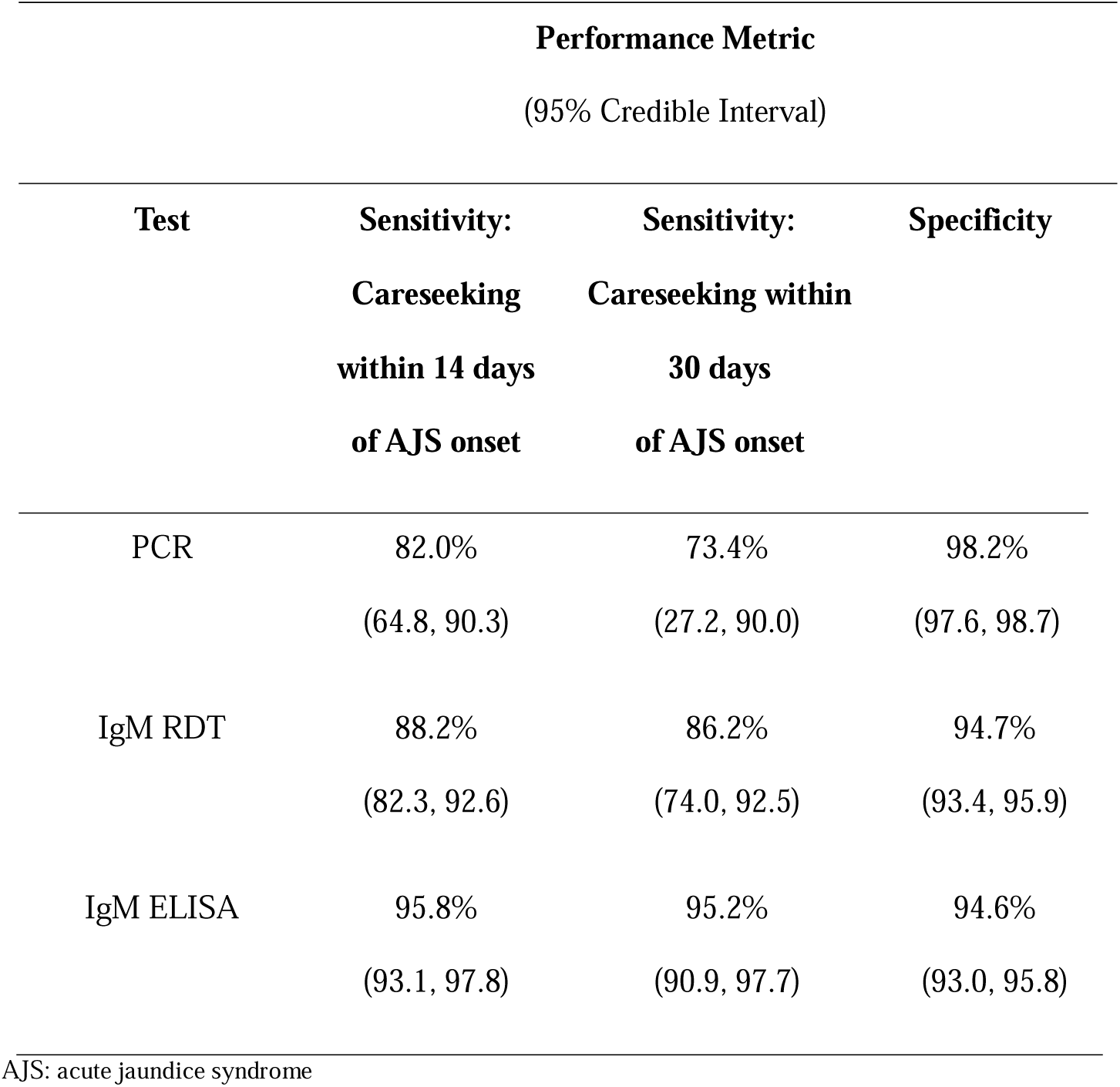
Sensitivity and specificity of diagnostic tests for detecting true hepatitis E infections based on latent class analysis.

| Performance Metric<br>(95% Credible Interval) |  |  |  |
| --- | --- | --- | --- |
| Test | Sensitivity:<br>Careseeking<br>within 14 days<br>of AJS onset | Sensitivity:<br>Careseeking within<br>30 days<br>of AJS onset | Specificity |
| PCR | 82.0%<br>(64.8, 90.3) | 73.4%<br>(27.2, 90.0) | 98.2%<br>(97.6, 98.7) |
| IgM RDT | 88.2%<br>(82.3, 92.6) | 86.2%<br>(74.0, 92.5) | 94.7%<br>(93.4, 95.9) |
| IgM ELISA | 95.8%<br>(93.1, 97.8) | 95.2%<br>(90.9, 97.7) | 94.6%<br>(93.0, 95.8) |

AJS: acute jaundice syndrome

Among suspected cases with careseeking delays up to 30 days the average positive predictive value (PPV) for PCR was 85.7% (95% CI: 37.0, 98.1), IgM RDT was 75.2% (95% CI: 20.2, 95.1), and IgM ELISA was 76.2% (95% CI: 21.3, 95.4). The average negative predictive value (NPV) among suspected cases with careseeking delays up to 30 days for PCR was 91.6% (95% CI: 67.0, 99.7), for IgM RDT was 95.0% (95% CI: 83.5, 99.8), and for IgM ELISA was 98.2% (95% CI: 93.5, 99.9).

As PPV and NPV are influenced by careseeking delays (through impacting senstivity) and the underlying prevalence of hepatitis E, we explored different hypothetical scenarios to demonstrate how test interpretation may differ in other settings. As prevalence increased, the PPV of all tests was higher and with less differences between tests (Supplemental Figure 2). Increasing careseeking delays and prevalence reduced the NPV of all tests but had the strongest effect on the NPV of PCR.

**Figure 2.**
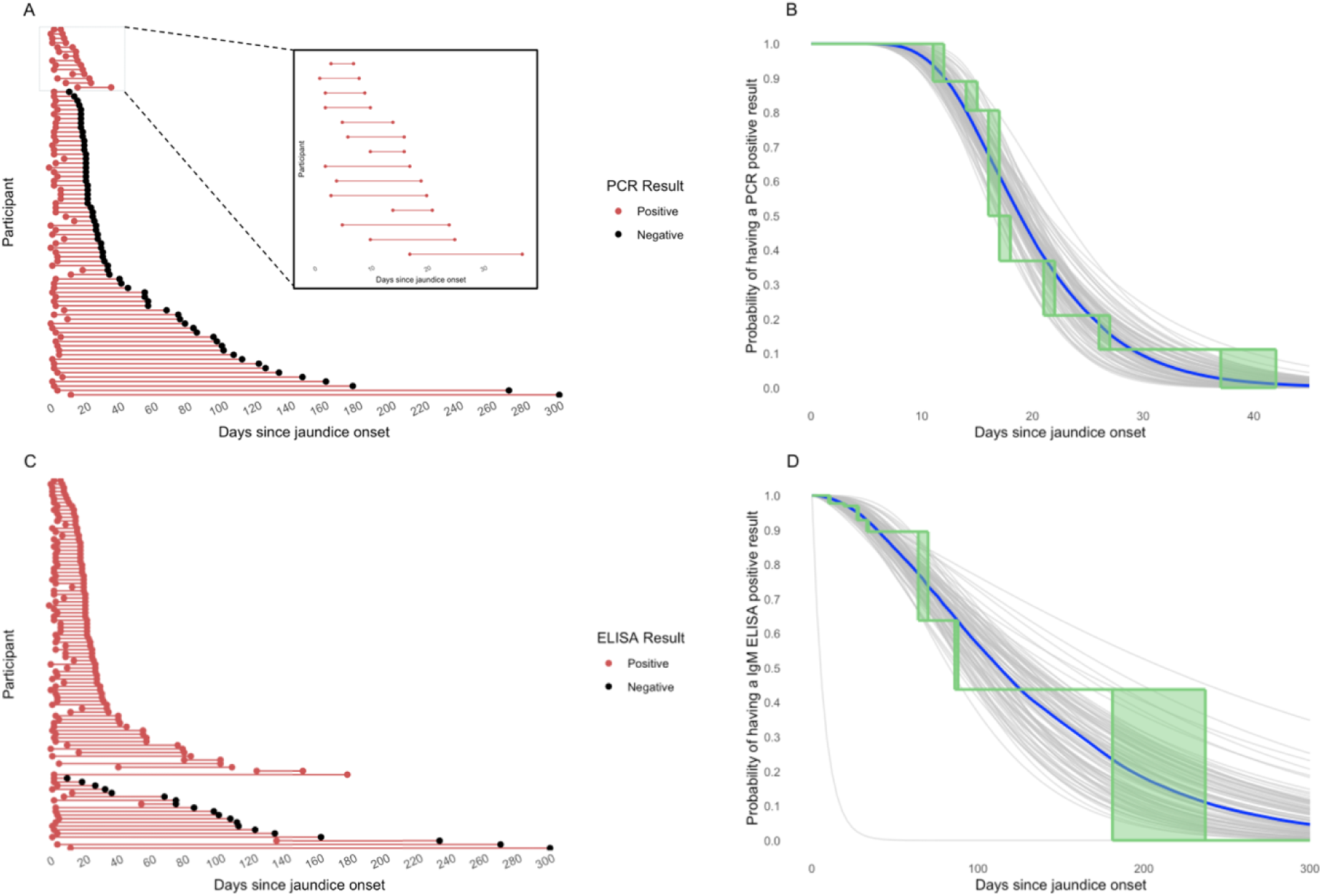
Longitudinal PCR and IgM ELISA results among suspected hepatitis E cases with a positive PCR and/or IgM ELISA result at enrollment. A: Cohort plot of participants who were PCR positive at enrollment with a follow-up test result. Each entry on the y-axis represents one participant. B: Parametric and non-parametric estimates of the probability of viremia at follow-up among participants who were viremic at enrollment based on serum PCR by days since jaundice onset. The best fitting parametric model assumed a log-normal distribution. C: Cohort plot of participants who were IgM ELISA positive at enrollment with a follow-up test result. D: Parametric and non-parametric estimates of the probability of IgM seropositivity at follow-up among participants who were seropositive based on serum ELISA at enrollment by days since jaundice onset. The best fitting parametric model assumed a gamma distribution. Rectangular regions in panel B and D represent estimates with similar likelihood in the non-parametric survival curve. Grey curves represent bootstrapped survival probability curves, blue curve represents median of all bootstrapped curves. Excludes 4 suspected cases who had indeterminate IgM ELISA results at follow-up.

### Time course of HEV viremia and IgM antibodies

In the subset of participants that returned for a follow-up visit with a blood draw (N=363), samples at enrollment were collected a median of 4 days after jaundice onset (IQR: 3-9; range: 0-217) while follow-up samples were collected a median of 36 days after jaundice onset (IQR: 21,77; range: 7-470) (Supplemental Table 7).

Among suspected cases who were PCR-positive at enrollment (N=83) only 14 (17%) were positive at follow-up (Figure 2), with a median time from jaundice onset to having a negative PCR result of 18.9 days (95% CI: 16.9, 21.3). Times to a negative PCR-result did not vary significantly by age or sex.

Among suspected cases who were IgM ELISA-positive at enrollment (N=104), 81 (78%) were still positive at follow-up, with a median time from jaundice onset to a negative IgM ELISA of 113.0 days (95% CI: 87.4, 163.1). Time to a negative IgM ELISA did not vary by sex but individuals under 5 years old had a faster IgM antibody decay (median: 61.6 days; 95% CI: 39.4, 90.2) compared to individuals over 5 (median: 125.8 days; 95% CI: 93.1, 184.5) (Supplemental Figure 7).

## Discussion

Diagnostic tests for hepatitis E had high sensitivity and specificity in field conditions, though sensitivity decreased with delayed careseeking post-jaundice onset. The Assure RDT demonstrated higher sensitivity than PCR and similar specificity to ELISA, underscoring its potential utility in surveillance for understanding burden of disease and detecting outbreaks. Negative PCR tests may not rule out current hepatitis E, particularly among individuals with modest delays between jaundice onset and careseeking. Our findings provide a pathway for strengthening diagnostic capacities for identifying hepatitis E cases in settings that are most likely to have limited laboratory infrastructure and be most impacted by hepatitis E genotypes 1 and 2.

Our crude estimates of IgM RDT sensitivity compared to PCR and/or IgM ELISA are similar to existing published literature from laboratory studies (11). Our estimates of PPV are not generalizable to other settings with different underlying hepatitis E prevalence and care seeking behavior, but our results can be used by decision-makers to generate setting-specific ranges of plausible PPVs. Though PCR/ELISA confirmation may still be necessary for official outbreak declarations, RDT results could help expedite outbreak response activities and help interrupt hepatitis E transmission in the critical early days of outbreaks.

Our adjusted estimates of the average sensitivity and specificity of each diagnostic test are based on the distribution of days between jaundice onset and care seeking in our study population. The proportion of suspected cases in our study with jaundice onset within 2 weeks of clinic visit at enrollment (82%) is notably higher than among hospitalized patients in an acute jaundice surveillance study in Bangladesh (39%) (25). Differences in careseeking could be due to the potentially shorter distance needed to travel to the MSF hospital within Bentiu IDP camp or due to increased care seeking given awareness about the ongoing HEV outbreak. Careseeking delays in our study population are similar to observed delays during a hepatitis E outbreak in a displaced persons camp in Darfur, Sudan (5 days) (26), suggesting careseeking delays may be shorter in camp settings with free healthcare compared to outside of camps for displaced persons. Notably, the PPV of RDTs did not vary greatly across different hypothetical delays in careseeking (Supplemental Figure 2).

We found faster IgM seroreversion in children under 5 compared to others, and some evidence HEV RNA clearance may also be faster in children under 5, though to a lesser extent. For some viruses, young children don’t mount robust antibody responses upon primary exposure and have lower peak antibody titers relative to older individuals (27). Lower peak antibody concentrations in young children, even if IgM decay rates were similar to adults, would lead to quicker IgM seroreversion. Further stratifying children in our study population illustrates that children under 4 years old had faster IgM decay compared to children 4 to 5 years (Supplemental Figure 8), suggesting that the youngest children are likely driving this trend. Interestingly, studies from Bangladesh have shown more frequent seroreversion of anti-HEV IgG antibodies in young children (28,29). Additional research tracking quantative measures of IgM antibodies can help better explain the apparent age-related differences in kinetics.

Our analyses have a number of limitations. Jaundice onset date in our study was self-reported, and jaundice can be difficult to identify in general and particularly in populations with darker skin (30,31). If self-reported jaundice onset date is systematically reported to be later than true date, we would underestimate the number of days between jaundice onset and care seeking. We expect this type of misclassification to have occurred non-differentially among HEV infected and uninfected suspected hepatitis E cases. Importantly, our results are based only on cases seeking care at the hospital (inpatient and outpatient). Our estimates for the sensitivity of each diagnostic test could be overestimates if cases with milder symptoms, lower HEV viral load, and/or lower IgM response were less likely to seek care. Our sensitivity and specificity estimates may not be generalizable to other settings due to Bentiu IDP camp’s history of repeated hepatitis E outbreaks. Infection with HEV generates long-lasting IgG antibodies, which could influence the sensitivity of diagnostic tests in subsequent infections. IgG positivity can reflect antibodies from a previous HEV infection, current HEV infection, or both. Animals previously exposed to HEV with high avidity anti-HEV IgG antibodies had shorter viremia, lower HEV RNA levels, and no detectable IgM antibody response compared to animals with lower avidity IgG when reinfected with HEV genotype 1 (32). We used venous blood for RDTs as opposed to capillary blood, which could impact RDT performance if venous blood has different fluid dynamics in lateral flow chromatographic assays and depending on staff training and comfort performing each procedure according to manufacturer’s instructions. Less than half of suspected cases returned for a follow-up visit.

Hepatitis E severity may be associated antibody response, therefore if individuals who returned for testing were less likely to have been severe cases, our estimates of median time from cases’ first ELISA positive result to having a negative result may be underestimated.

We provide new insights on both the time-varying sensitivity of IgM RDTs and the traditional ‘gold-standard’ assays, which can help inform test interpretation and future surveillance guidance. Strengthening investments in surveillance in settings where hepatitis E transmission occurs can help us more rapidly detect and respond to outbreaks and develop more data-informed strategies to using hepatitis E vaccines both in endemic and epidemic settings.

## Data Availability

Code for all analyses and a minimal dataset are available at https://github.com/HopkinsIDD/hev-diagnostics-bentiu.

## Acknowledgements

We thank all the MSF staff in Bentiu who collected these data and the participants in Bentiu IDP camp. We thank the diagnostic laboratory of the University Hospitals of Geneva for help with serological testing. We thank Javier Perez-Saez and Benjamin Meyer for the helpful discussions and technical inputs.

## Funding

This work was supported by Médecins Sans Frontières. The funders of the study were fully involved in the study design, data collection, data analysis, data interpretation and writing of the report.

## Conflicts of Interest

The authors have no conflicts of interest to declare.

## Author contributions

Conceptualization: AK, ASA, IC, RN; Methodology: AK, ASA; Formal analysis: AK, CA; Investigation: KVA, RN, CA; Resources: KVA, RN, IE; Data curation: KVA, CA; Writing – Original Draft: AK; Writing – Review & Editing: CA, EG, JW, MH, MA, EG, ESG, FBL, DB, MR, JR, IC, IE, ASA; Supervision: ASA, RN, IE; Project administration: RN, KVA, CA, EG, MA

## Notes

### Competing Interest Statement

The authors have declared no competing interest.

### Funding Statement

This work was supported by Medecins Sans Frontieres. The funders of the study were fully involved in the study design, data collection, data analysis, data interpretation and writing of the report.

### Author Declarations

Ethical approval for the parent study was obtained from Medecins Sans Frontiere (ERB #2167), the South Sudan Ministry of Health Research Ethics Board (RERB-MOH # 54/27/09/2022) and The Johns Hopkins Bloomberg School of Public Health Institutional Review Board (IRB00025966).

