## Supplemental Figures and Tables for "Diagnostic performance and kinetics of hepatitis E viral RNA and IgM antibody test positivity in a genotype 1 outbreak in South Sudan"

**Supplement**

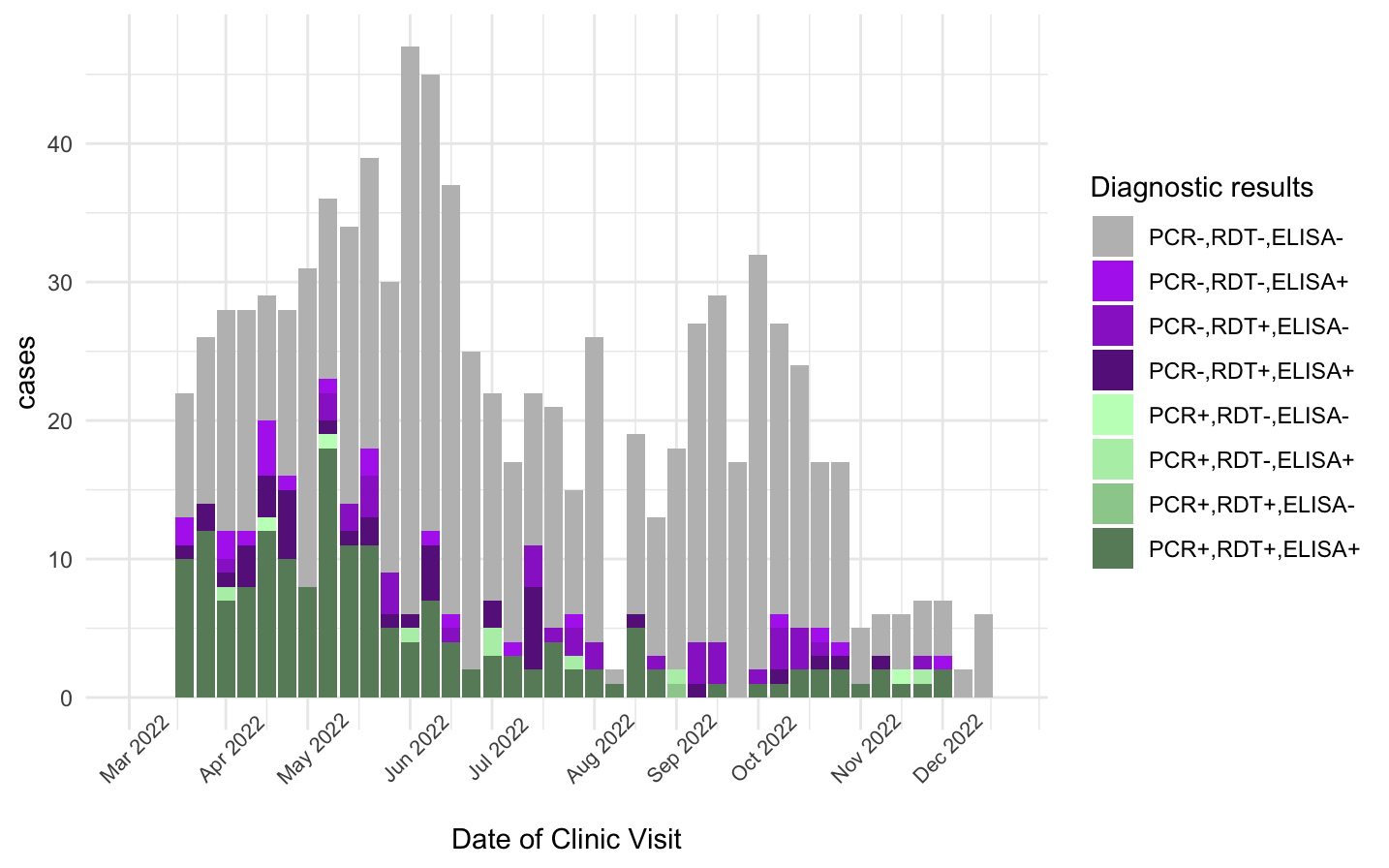

**Supplemental Figure 1. Epidemic curve for suspected hepatitis E cases stratified by diagnostic test results, Bentiu, 2022**

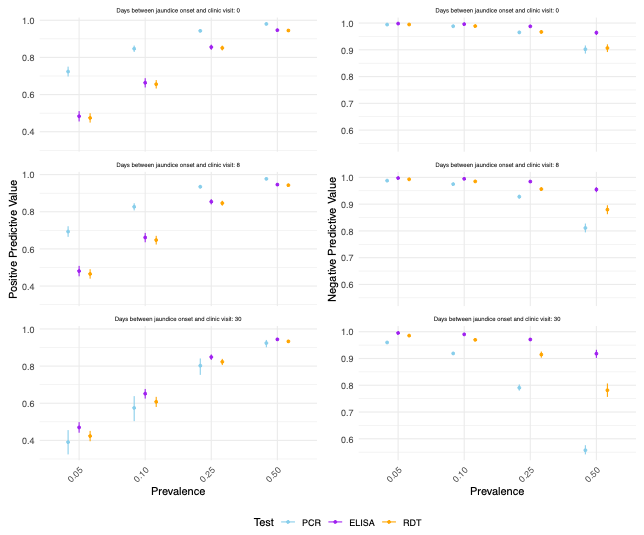

**Supplemental Figure 2. Positive and negative predictive value of diagnostic tests for detecting true hepatitis E infections by different prevalence and careseeking scenarios.** We used posterior estimates of the following parameters to estimate the positive and negative predictive values of each test: intercepts and spline coefficients in each regression of sensitivity on days between self-reported jaundice onset and clinic visit, specificity, and time-varying risk of hepatitis E. Points indicate median and error bars indicate 20^th^ and 80^th^ quantiles across all posterior draws. Days between jaundice onset and clinic visit of 8 days represents the median careseeking behavior of our study population including enrollment and follow-up visits.

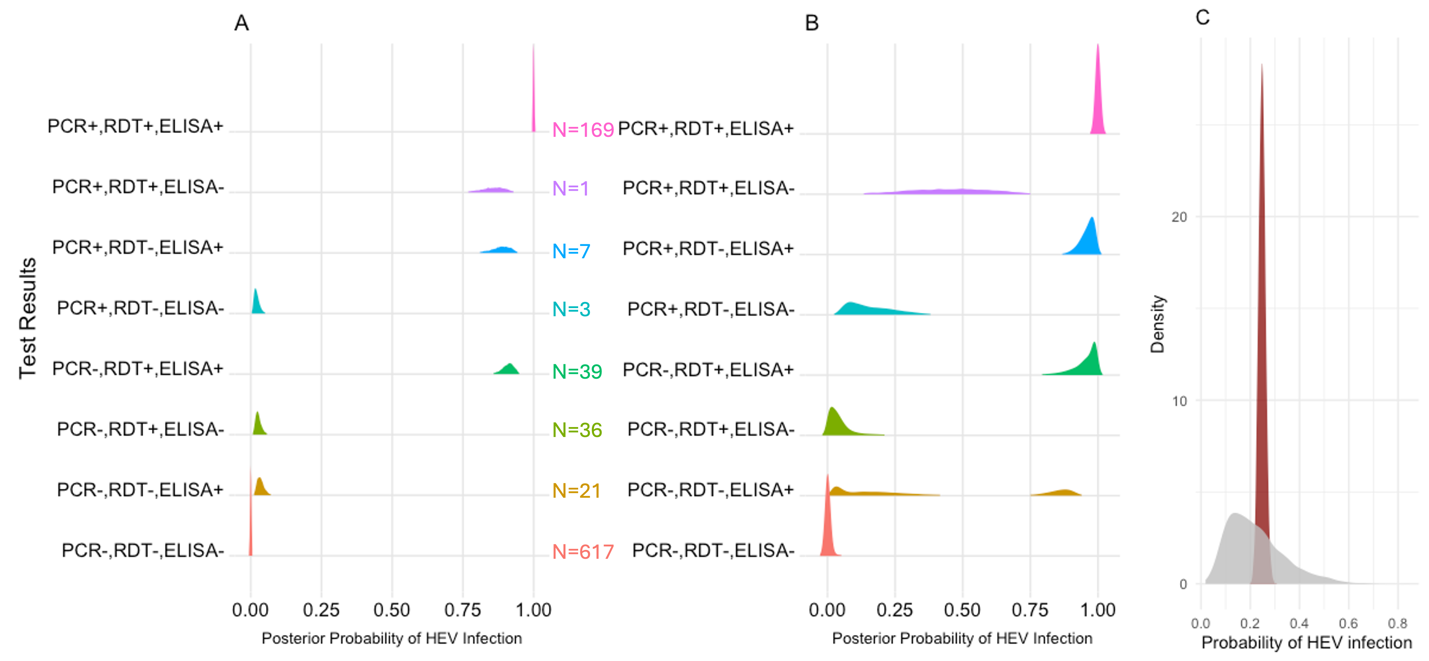

**Supplemental Figure 3. Predicted probability of hepatitis E virus (HEV) infection stratified by test results at enrollment in the unadjusted latent class model (A), stratified by test results at enrollment in the adjusted latent class model allowing sensitivity to decline as a linear function of days between jaundice onset and clinic visit (B), and in the full sample in the adjusted latent class model (C).** Grey shading in panel C indicates the prior predictive distribution and maroon indicates the posterior distribution. Sample size by test result was the same in the unadjusted and adjusted latent class models.

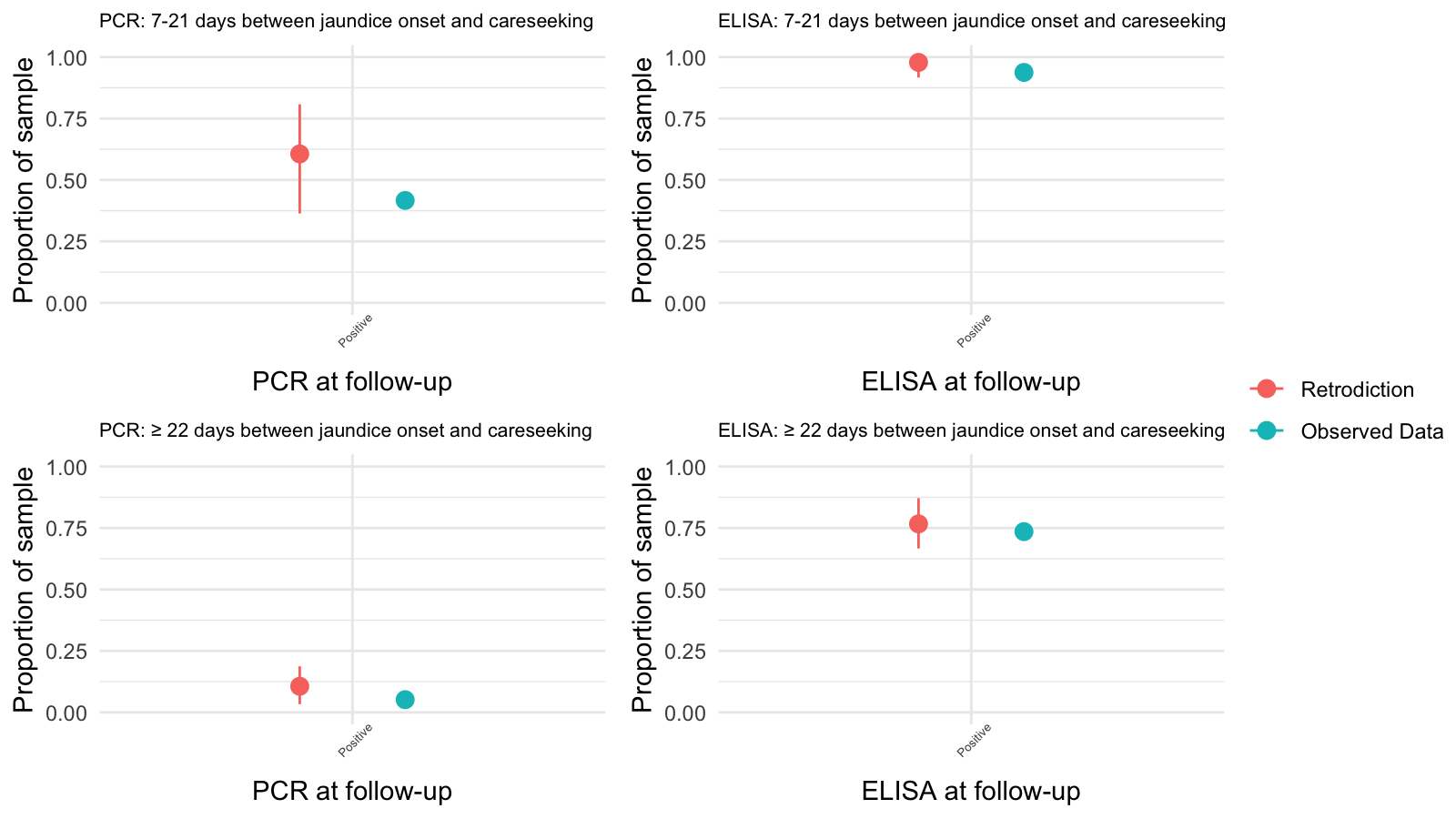

**Supplemental Figure 4. Posterior retrodictive checks of follow-up PCR and IgM ELISA test results among individuals who were PCR and/or IgM ELISA positive at enrollment in observed data compared to the test results predicted by accelerated failure time models, stratified by days between jaundice onset and careseeking.** Error bars indicate 2·5^th^ and 97·5^th^ quantiles from bootstrapping.

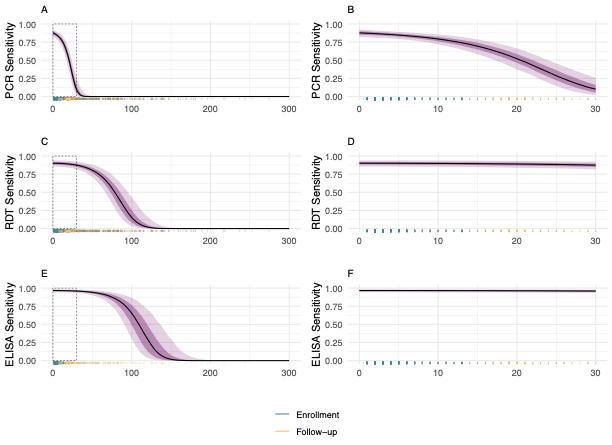

**Supplemental Figure 5. Sensitivity and specificity of diagnostic tests for detecting hepatitis E infections as a function of days between jaundice onset and clinic visit using cubic splines.** A: PCR sensitivity across all careseeking delays with box around the first 30 days between jaundice onset and clinic visit; B: PCR sensitivity in the first 30 days between jaundice onset and clinic visit; C: IgM RDT sensitivity across all careseeking delays with box around the first 30 days between jaundice onset and clinic visit; D: IgM RDT in the first 30 days between jaundice onset and clinic visit; E: IgM ELISA sensitivity across all careseeking delays with box around the first 30 days between jaundice onset and clinic visit; F: IgM ELISA in the first 30 days between jaundice onset and clinic visit; Rug plot showing frequency of observed values. Line indicates median, dark purple indicates 20^th^ and 80^th^ quantiles, and light purple indicates 2·5^th^ and 97·5^th^ quantiles. Logit sensitivity varied by days between self-reported jaundice onset date and clinic visit using a cubic spline with three degrees of freedom, separately for each test.

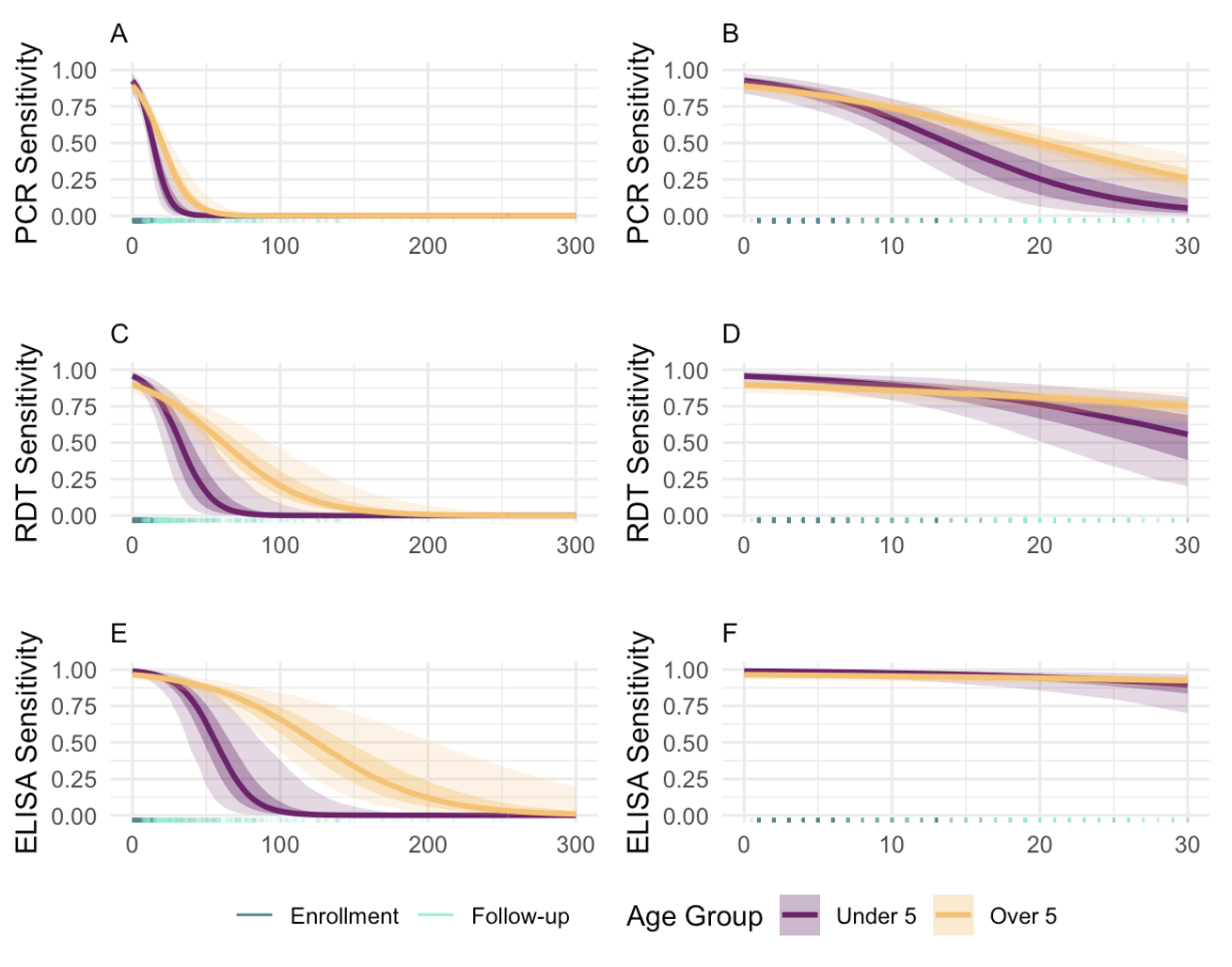

**Supplemental Figure 6. Sensitivity and specificity of diagnostic tests for detecting hepatitis E infections as a linear function of days between jaundice onset and clinic visit, allowing decline in sensitivity to vary by age group.** A: PCR sensitivity across all careseeking delays with box around the first 30 days between jaundice onset and clinic visit; B: PCR sensitivity in the first 30 days between jaundice onset and clinic visit; C: IgM RDT sensitivity across all careseeking delays with box around the first 30 days between jaundice onset and clinic visit; D: IgM RDT in the first 30 days between jaundice onset and clinic visit; E: IgM ELISA sensitivity across all careseeking delays with box around the first 30 days between jaundice onset and clinic visit; F: IgM ELISA in the first 30 days between jaundice onset and clinic visit; Rug plot showing frequency of observed values. Line indicates median, darker color around median indicates 20^th^ and 80^th^ quantiles, and lighter color indicates 2·5^th^ and 97·5^th^ quantiles. Logit sensitivity varied linearly by days between self-reported jaundice onset and clinic visit, separately for each test.

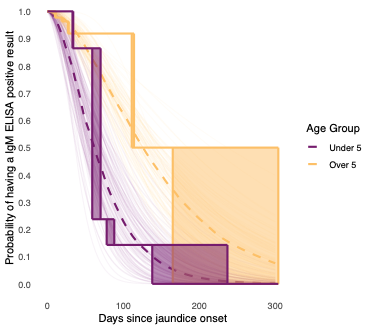

**Supplemental Figure 7. Parametric and non-parametric estimates of the probability of IgM ELISA seropositivity at follow-up by days since jaundice onset, stratified by age under and over 5.** The best fitting parametric model assumed a gamma distribution. Rectangular regions represent estimates with similar likelihood in the non-parametric survival curve. Curves represent bootstrapped survival probability curves by age group, dashed curve represents median of all bootstrapped curves by age group. Excludes four suspected cases who had indeterminate IgM ELISA results at follow-up.

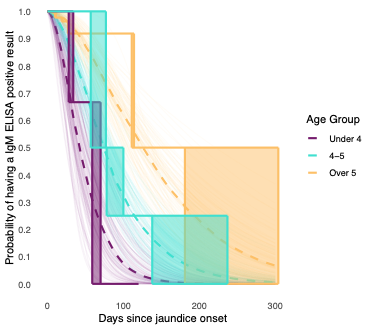

**Supplemental Figure 8. Parametric and non-parametric estimates of the probability of IgM ELISA seropositivity at follow-up by days since jaundice onset, stratified by age under 4, age 4-5, and age over 5.**

The best fitting parametric model assumed a gamma distribution. Rectangular regions represent estimates with similar likelihood in the non-parametric survival curve. Curves represent bootstrapped survival probability curves by age group, dashed curve represents median of all bootstrapped curves by age group. Excludes four suspected cases who had indeterminate IgM ELISA results at follow-up.

**Supplemental Table 1.** **Characteristics of suspected hepatitis E cases who presented to care at a health facility between March and December 2022**

|  | | **Case type** | | | | | | | |
| --- | --- | --- | --- | --- | --- | --- | --- | --- | --- |
| **Characteristic**  **N (col %)** | Overall N=893 | PCR+  RDT+ ELISA+  N=169 | PCR+ RDT+ ELISA-  N=1 | PCR+  RDT- ELISA+  N=7 | PCR+  RDT- ELISA-  N=3 | PCR- RDT+ ELISA+  N=39 | PCR-  RDT+  ELISA-  N=36 | PCR- RDT- ELISA+  N=21 | PCR- RDT- ELISA-  N=617 |
| Female | 406 (45·5) | 77 (45·6) | 1 (100) | 3 (42·9) | 2 (66·7) | 17 (43·6) | 16 (44·4) | 9 (42·9) | 281 (45·5) |
| Age, median (IQR) | 15·5  (5·9, 26·2) | 9·6  (4·9, 17·7) | 38·3  (38·3, 38·3) | 10·5  (5·8, 19·8) | 3·9  (2·6, 7·3) | 10·2  (4·1, 17·2) | 22·1  (11·7, 32·3) | 8·1  (5·5, 14·8) | 18·3  (6·9, 28·5) |
| Age |  |  |  |  |  |  |  |  |  |
| 0-5 | 192 (21·5) | 44 (26·0) | 0 (0) | 2 (28·6) | 2 (66·7) | 12 (30·8) | 5 (13·9) | 5 (23·8) | 122 (19·8) |
| 6-15 | 241 (27·0) | 69 (40·8) | 0 (0) | 2 (28·6) | 1 (33·3) | 14 (35·9) | 6 (16·7) | 11 (52·4) | 138 (22·4) |
| 16-39 | 379 (42·4) | 50 (29·6) | 1 (100) | 3 (42·9) | 0 (0) | 11 (28·2) | 18 (50·0) | 5 (23·8) | 291 (47·2) |
| 40+ | 81 (9·1) | 6 (3·6) | 0 (0) | 0 (0) | 0 (0) | 2 (5·1) | 7 (19·4) | 0 (0) | 66 (10·7) |
| Days since jaundice onset*, median (IQR) | 5·0  (3·0, 10·0) | 4·0  (3·0, 7·0) | 5·0  (5·0, 5·0) | 3·0  (3·0, 5·0) | 11·0  (6·5, 14·0) | 5·0  (3·0, 17·5) | 5·0  (3·0, 11·5) | 6·0  (3·0, 51·0) | 5·0  (3·0, 12·0) |
| Days since jaundice onset* |  |  |  |  |  |  |  |  |  |
| ≤1 week | 602 (67·4) | 132 (78·1) | 1 (100) | 6 (85·7) | 1 (33·3) | 23 (59·0) | 22 (61·1) | 11 (52·4) | 406 (65·9) |
| >1 week to 2 weeks | 129 (14·4) | 30 (17·8) | 0 (0) | 0 (0) | 1 (33·3) | 5 (12·8) | 6 (16·7) | 3 (14·3) | 84 (13·6) |
| >2 weeks to 1 month | 74 (8·3) | 6 (3·6) | 0 (0) | 1 (14·3) | 1 (33·3) | 4 (10·3) | 4 (11·1) | 0 (0) | 58 (9·4) |
| >1 month to 2 months | 49 (5·5) | 1 (0·6) | 0 (0) | 0 (0) | 0 (0) | 4 (10·3) | 2 (5·6) | 3 (14·3) | 39 (6·3) |
| >2 months | 38 (4·3) | 0 (0) | 0 (0) | 0 (0) | 0 (0) | 3 (7·7) | 2 (5·6) | 4 (19·0) | 29 (4·7) |
| Anti-HEV IgG positive | 763 (85·4) | 169 (100) | 1 (100) | 6 (85·7) | 2 (66·7) | 39 (100) | 30 (83·3) | 19 (90·5) | 497 (80·6) |
| Elevated ALT** | 135 (20·5) | 98 (72·1) | 0 (0) | 1 (16·7) | 0 (0) | 2 (6·9) | 1 (4·5) | 0 (0) | 33 (7·4) |
| Elevated AST*** | 55 (17·5) | 8 (25·0) | 0 (0) | 0 (0) | 0 (0) | 2 (18·2) | 2 (11·1) | 0 (0) | 43 (17·6) |
| Hospitalized**** | 57 (6·4) | 6 (3·6) | 1 (100) | 0 (0) | 0 (0) | 5 (12·8) | 7 (19·4) | 0 (0) | 38 (6·2) |
| Died | 9 (1·0) | 0 (0) | 0 (0) | 0 (0) | 0 (0) | 1 (2·6) | 1 (2·8) | 0 (0) | 7 (1·1) |
| Attended follow-up visit | 363 (40·6) | 77 (45·6) | 0 (0) | 5 (71·4) | 1 (33·3) | 15 (38·5) | 15 (41·7) | 7 (33·3) | 243 (39·4) |

*Missing for 1 suspected cases

**Missing for 233 suspected cases. For males, ALT ≥41 units per liter was considered elevated. For females, ALT >32 units per liter was considered elevated (23,24).

***Missing for 578 suspected cases. AST above 40 units per liter was considered elevated.

****Missing for 1 suspected case

**Supplemental Table 2. Observed test results and sensitivity and specificity compared to polymerase chain reaction (PCR) and IgM enzyme-linked immunosorbent assay (ELISA)**

|  | **Gold standard** | | **Metric** (95% CI) | |
| --- | --- | --- | --- | --- |
|  | **PCR+ or**  **IgM ELISA+** | **PCR- and IgM ELISA-** | **Sensitivity** | **Specificity** |
| **IgM RDT+** | 209 | 36 | 87·1%  (82·2, 91·1) | 94·5%  (92·4, 96·1) |
| **IgM RDT-** | 31 | 616 |  |  |
|  | **PCR+ and IgM ELISA+** | **PCR- and IgM ELISA-** | **Sensitivity** | **Specificity** |
| **IgM RDT+** | 169 | 36 | 96·0%  (92·0, 98·4) | 94·5%  (92·4, 96·1) |
| **IgM RDT-** | 7 | 616 |  |  |
| **Test results** | **PCR+** | **PCR-** | **Sensitivity** | **Specificity** |
| **IgM ELISA+** | 176 | 60 | 97·8%  (94·4, 99·4) | 91·6%  (89·3, 93·5) |
| **IgM ELISA-** | 4 | 652 |  |  |

**Supplemental Table 3. Sensitivity analyses comparing latent class models varying the functional form of days since jaundice onset on sensitivity of PCR, IgM RDT, and IgM ELISA and adjusting for age and sex.** Grey shading indicates final selected model.

| **Days since**  **jaundice onset*** | | | **Time Varying**  **Risk of Hepatitis E**** | **Age*** | **Interaction*: Age and days since jaundice onset** | **Sex*** | **Sensitivity:**  **Careseeking within 14 days of AJS onset** | | | **Specificity** | | |
| --- | --- | --- | --- | --- | --- | --- | --- | --- | --- | --- | --- | --- |
| **PCR** | **IgM RDT** | **IgM ELISA** |  |  |  |  | **PCR** | **IgM RDT** | **IgM ELISA** | **PCR** | **IgM RDT** | **IgM ELISA** |
| Linear | Linear | Linear |  |  |  |  | 83·3% (66·1, 90·9) | 88·1%  (92·8, 92·1) | 93·7%  (90·8, 96·0) | 98·2%  (97·6, 98·7) | 94·8%  (93·5, 95·9) | 94·4%  (92·9, 95·9) |
| Linear | Linear | Linear | X |  |  |  | 82·0%  (64·8, 90·3) | 88·2% (82·3, 92·6) | 95·8%  (93·1, 97·8) | 98·2%  (97·6, 98·7) | 94·7%  (93·4, 95·9) | 94·6%  (93·0, 95·8) |
| Linear | Linear | Linear | X | X | X |  | 82·4%  (64·5, 92·2) | 88·9%  (81·5, 96·7) | 96·3%  (92·8, 99·4) | 98·2%  (97·6, 98·7) | 94·7%  (93·4, 95·9) | 94·5%  (92·9, 95·8) |
| Linear | Linear | Linear | X | X |  |  | 82·1%  (64·7, 90·9) | 88·2%  (81·1, 94·8) | 95·9%  (92·5, 98·5) | 98·2%  (97·6, 98·7) | 94·7%  (93·4, 95·9) | 94·5%  (93·0, 95·9) |
| Linear^¶^ | Linear^¶^ | Linear^¶^ | X |  |  |  | 83·6%  (72·5, 90·3) | 89·7%  (84·4, 93·9) | 95·8%  (93·1, 97·7) | 98·2%  (97·6, 98·7) | 94·7% (93·4, 95·9) | 94·1%  (92·5, 95·5) |
| Linear | Linear | Linear |  |  |  | X | 82·3%  (65·5, 90·3) | 89·2%  (83·6, 93·4) | 96·6%  (94·2, 98·3) | 98·2%  (97·6, 98·7) | 94·7%  (93·5, 95·9) | 94·3%  (92·7, 95·7) |
| Linear | Linear | - | X |  |  |  | 82·4%  (65·8, 90·4) | 89·1%  (83·5, 93·3) | 96·8%  (94·3, 98·4) | 98·2%  (97·6, 98·7) | 94·7%  (93·4, 95·8) | 94·6%  (93·1, 95·9) |
| - | Linear | Linear |  |  |  |  | 79·1%  (72·9, 85·0) | 93·5%  (89·5, 96·5) | 97·0%  (94·9, 98·5) | 98·2%  (97·6, 98·7) | 94·7%  (93·5, 95·9) | 92·1%  (90·4, 93·7) |
| - | Linear | Linear | X |  |  |  | 76·8%  (70·1, 82·9) | 92·4%  (88·0, 95·9) | 97·0%  (94·8, 98·5) | 98·2%  (97·6, 98·7) | 94·7%  (93·4, 95·8) | 92·6%  (90·9, 94·2) |
| Linear | - | Linear | X |  |  |  | 82·9%  (67·4, 90·6) | 90·0%  (84·5, 94·3) | 96·7%  (94·4, 98·4) | 98·2%  (97·6, 98·7) | 94·8%  (93·5, 95·9) | 93·8%  (92·1, 95·2) |
| Linear | - | - |  |  |  |  | 83·0%  (68·0, 90·5) | 91·5%  (86·7, 95·4) | 96·9%  (94·7, 98·5) | 98·2%  (97·6, 98·7) | 94·8%  (93·4, 95·9) | 93·5%  (91·8, 95·0) |
| Linear | - | - | X |  |  |  | 83·0%  (67·5, 90·7) | 90·3%  (85·1, 94·5) | 97·0%  (94·9, 98·5) | 98·2%  (97·6, 98·7) | 94·7%  (93·5, 95·9) | 93·8%  (92·2, 95·2) |
| Cubic  spline | Cubic  spline | Cubic  spline |  |  |  |  | 84·1%  (72·7, 90·3) | 90·7%  (85·8, 94·6) | 96·7%  (94·3, 98·3) | 98·2%  (97·6, 98·7) | 94·7%  (93·5, 95·9) | 93·9%  (92·4, 95·4) |
| Cubic  spline | Cubic  spline | Cubic  spline | X |  |  |  | 84·0%  (72·5, 90·3) | 89·8%  (84·9, 93·9) | 96·5%  (94·2, 98·2) | 98·2%  (97·6, 98·7) | 94·7%  (93·3, 95·8) | 94·0%  (92·4, 95·5) |
| Cubic  spline | Cubic  spline | - | X |  |  |  | 84·1%  (72·6, 90·2) | 90·3%  (85·3, 94·2) | 96·8%  (94·4, 98·5) | 98·2%  (97·6, 98·7) | 94·7%  (93·4, 95·8) | 94·2%  (92·7, 95·6) |
| Cubic  spline | Linear | - | X |  |  |  | 83·8%  (71·7, 90·2) | 89·1%  (83·6, 93·3) | 96·8%  (94·3, 98·5) | 98·2%  (97·6, 98·7) | 94·7%  (93·4, 95·8) | 94·6%  (93·2, 95·8) |
| Cubic  spline | Linear | Linear |  |  |  |  | 83·5%  (71·1, 90·0) | 89·1%  (83·5, 93·3) | 96·2%  (93·6, 98·0) | 98·2%  (97·6, 98·7) | 94·8%  (93·5, 95·9) | 94·5%  (92·9, 95·8) |
| Cubic  spline | - | - |  |  |  |  | 84·3%  (73·7, 90·2) | 91·4%  (86·4, 95·3) | 96·9%  (94·6, 98·5) | 98·2%  (97·6, 98·7) | 94·8%  (93·5, 95·9) | 93·6%  (91·9, 95·1) |
| Cubic  spline | - | - | X |  |  |  | 84·4%  (73·4, 90·5) | 90·3%  (85·2, 94·5) | 97·0%  (94·8, 98·6) | 98·2% (97·6, 98·7) | 94·7%  (93·4, 95·9) | 93·8%  (92·2, 95·3) |
| - | Cubic  spline | Cubic  spline |  |  |  |  | 78·7%  (72·4, 84·5) | 93·8%  (90·2, 96·6) | 97·0%  (94·9, 98·6) | 98·2%  (97·7, 98·7) | 94·8%  (93·4, 95·9) | 92·1%  (90·4, 93·6) |
| - | Cubic  spline | Cubic  spline | X |  |  |  | 76·6%  (70·3, 82·6) | 93·1%  (89·1, 96·4) | 97·1%  (95·1, 98·6) | 98·2%  (97·6, 98·7) | 94·7%  (93·5, 95·8) | 92·5%  (90·8, 94·0) |
| Linear | Cubic  spline | Linear |  |  |  |  | 82·7%  (66·8, 90·4) | 90·6%  (85·9, 94·5) | 96·3%  (93·9, 98·1) | 98·2%  (97·6, 98·7) | 94·8%  (93·6, 95·9) | 94·0%  (92·5, 95·4) |
| Linear | Linear | Cubic  spline |  |  |  |  | 82·3%  (65·5, 90·3) | 89·2%  (83·6, 93·4) | 96·6%  (94·2, 98·3) | 98·2%  (97·6, 98·7) | 94·7%  (93·5, 95·9) | 94·3%  (92·7, 95·7) |

AJS: acute jaundice syndrome

*Only included in models estimating logit sensitivity. All functional forms for days since jaundice onset are on the logit scale.

**Varied as a function of month of clinic visit using binary indicator variables for month

^¶^Adjusting for days since symptom onset in place of days since jaundice onset

**Supplemental Table 4. Model comparison using leave-one-out cross-validation**. Difference in ELPD is a relative difference between the expected log predictive density (ELPD) of all models compared to the model with the largest ELPD. The difference in standard error (SE) represents the standard error of the difference in ELPD between pairs of models. Grey shading indicates final selected model.

| **Days since jaundice onset*** | | | **Time**  **Varying**  **Risk of Hepatitis E**** | **Age*** | **Interaction*:**  **Age and days since jaundice onset** | **Sex*** | **Model comparison** | |
| --- | --- | --- | --- | --- | --- | --- | --- | --- |
| **PCR** | **IgM RDT** | **IgM ELISA** |  |  |  |  | Difference  in ELPD | Difference in SE |
| Linear | Linear | Linear | X | X | X |  | 0 | 0 |
| Linear | Linear | Linear | X |  |  |  | -0·3 | 4·1 |
| Linear | Linear | Linear | X | X |  |  | -2·2 | 4·1 |
| Cubic spline | Cubic spline | Cubic spline | X |  |  |  | -10·6 | 5·4 |
| Cubic spline | Linear | - | X |  |  |  | -17·1 | 6·3 |
| Linear | Linear | - | X |  |  |  | -17·7 | 5·8 |
| Cubic spline | Cubic spline | - | X |  |  |  | -23·3 | 6·6 |
| Linear | - | Linear | X |  |  |  | -35·2 | 6·4 |
| Cubic spline | - | - | X |  |  |  | -36·9 | 6·7 |
| Linear | - | - | X |  |  |  | -37·3 | 6·4 |
| Cubic spline | Linear | Linear |  |  |  |  | -61·9 | 11·6 |
| Linear | Linear | Linear |  |  |  | X | -61·9 | 11·3 |
| Linear | Linear | Cubic spline |  |  |  |  | -62·8 | 11·3 |
| Linear | Linear | Linear |  |  |  |  | -65·1 | 11·4 |
| - | Cubic spline | Cubic spline | X |  |  |  | -66·7 | 8·8 |
| - | Linear | Linear | X |  |  |  | -68·3 | 8·8 |
| Cubic spline | Cubic spline | Cubic spline |  |  |  |  | -68·5 | 11·6 |
| Linear | Cubic spline | Linear |  |  |  |  | -69·3 | 11·4 |
| - | - | - | X |  |  |  | -71·8 | 9·2 |
| Cubic spline | - | - |  |  |  |  | -85·0 | 12·1 |
| Linear | - | - |  |  |  |  | -85·7 | 11·9 |
| - | Cubic spline | Cubic spline |  |  |  |  | -112·7 | 13·6 |
| - | Linear | Linear |  |  |  |  | -114·0 | 13·7 |
| - | - | - |  | X |  | X | -114·4 | 13·8 |
| - | - | - |  |  |  |  | -115·2 | 13·7 |

*Only included in models estimating sensitivity

**Varied as a function of month of clinic visit using binary indicator variables for month

ELPD: expected log pointwise predictive density; SE: standard error; PCR: polymerase chain reaction; IgM RDT: immunoglobulin M rapid diagnostic test; IgM ELISA: immunoglobulin M enzyme-linked immunosorbent assay

**Supplemental Table 5. Prior predictive distributions for the final adjusted latent class model.** Final model adjusted for days since jaundice onset for each test using a cubic spline with three degrees of freedom.

| **Parameter*** | **Prior** |
| --- | --- |
| Risk of hepatitis E | Normal (-1·39, 0·7) |
| Coefficient for month of clinic visit (indicator variables) | Normal (0, 2) |
| Logit Sensitivity: PCR** | Normal (1·5, 0·5) |
| Logit Sensitivity: IgM RDT** | Normal (2·2, 0·5) |
| Logit Sensitivity: IgM ELISA** | Normal (2·3, 0·5) |
| Logit Specificity: PCR | Normal (3·5, 0·2) |
| Logit Specificity: IgM RDT | Normal (2·5, 0·2) |
| Logit Specificity: IgM ELISA | Normal (2·5, 0·2) |
| Coefficients for days since jaundice onset cubic spline: PCR | Spline term 1: Half-Normal (0, 0·25)  Spline term 2: Half-Normal (0, 0·001)  Spline term 3: Half-Normal (0, 0·001) |
| Coefficients for days since jaundice onset cubic spline: IgM RDT | Spline term 1: Half-Normal (0, 0·001)  Spline term 2: Half-Normal (0, 0·0001)  Spline term 3: Half-Normal (0, 0·000001) |
| Coefficients for days since jaundice onset cubic spline: IgM ELISA | Spline term 1: Half-Normal (0, 0·001)  Spline term 2: Half-Normal (0, 0·0001)  Spline term 3: Half-Normal (0, 0·000001) |

*Prior for the logit of all parameters

**Prior for intercept in regression of sensitivity on days between self-reported jaundice onset and clinic visit. Intercept represents the baseline sensitivity among suspected cases with 0 days between jaundice onset and clinic visit.

**Supplemental Table 6. Sensitivity analyses comparing final latent class model varying prior distributions for the sensitivity and specificity of PCR, RDT, and ELISA.** Grey shading indicates final selected model.

|  | | Priors | | | Performance (95% CI) | | | | | |
| --- | --- | --- | --- | --- | --- | --- | --- | --- | --- | --- |
|  |  |  |  |  | Sensitivity:  Careseeking within 14 days of AJS onset | | | Specificity | | |
|  |  | PCR | IgM RDT | IgM ELISA | PCR | IgM RDT | IgM ELISA | PCR | IgM RDT | IgM ELISA |
| A | Logit Sensitivity* | Normal  (1·5, 0·5) | Normal  (2·2, 0·5) | Normal  (2·3, 0·5) | 82·0%  (64·8, 90·3) | 88·2% (82·3, 92·6) | 95·8%  (93·1, 97·8) | 98·2%  (97·6, 98·7) | 94·7%  (93·4, 95·9) | 94·6%  (93·0, 95·8) |
|  | Logit Specificity | Normal  (3·5, 0·2) | Normal  (2·5, 0·2) | Normal  (2·5, 0·2) |  |  |  |  |  |  |
| B | Logit Sensitivity* | Normal  (1·5, 0·5) | Normal  (2·2, 0·5) | Normal  (1·99, 0·5) | 78·7%  (58·8, 88·2) | 84·3%  (77·4, 89·5) | 95·5%  (92·8, 97·5) | 98·2%  (97·6, 98·7) | 94·8%  (93·5, 96·0) | 98·8%  (98·3, 99·2) |
|  | Logit Specificity | Normal  (3·5, 0·2) | Normal  (2·5, 0·2) | Normal  (4·59,0·2) |  |  |  |  |  |  |
| C | Logit Sensitivity* | Normal  (1·5, 0·5) | Normal  (2·59,0·5) | Normal  (2·3, 0·5) | 80·6%  (63·3, 89·1) | 89·1%  (83·3, 93·5) | 94·8%  (91·3, 97·2) | 98·2%  (97·6, 98·7) | 98·2%  (97·6, 98·8) | 94·6%  (93·2, 95·9) |
|  | Logit Specificity | Normal  (3·5, 0·2) | Normal  (4·59,0·2) | Normal  (2·5, 0·2) |  |  |  |  |  |  |
| D | Logit Sensitivity* | Normal  (2·3, 0·5) | Normal  (2·2, 0·5) | Normal  (2·3, 0·5) | 83·6%  (66·1, 91·5) | 88·3%  (82·4, 92·7) | 95·9%  (93·2, 97·9) | 96·9%  (96·0, 97·7) | 94·7%  (93·4, 95·8) | 94·5%  (92·9, 95·8) |
|  | Logit Specificity | Normal  (2·5, 0·2) | Normal  (2·5, 0·2) | Normal  (2·5, 0·2) |  |  |  |  |  |  |
| E | Logit Sensitivity* | Normal  (1·99, 0·5) | Normal  (2·59,0·5) | Normal  (1·99, 0·5) | 78·5%  (58·3, 88·1) | 85·2%  (78·5, 90·2) | 94·1%  (90·7, 96·6) | 99·2%  (98·8, 99·4) | 98·3%  (97·6, 98·8) | 98·8%  (98·3, 99·2) |
|  | Logit  Specificity | Normal  (4·59,0·2) | Normal  (4·59,0·2) | Normal  (4·59,0·2) |  |  |  |  |  |  |

*Prior for intercept in regression of logit sensitivity on days between self-reported jaundice onset and clinic visit. Intercept represents the baseline logit sensitivity among suspected cases with 0 days between jaundice onset and clinic visit.

AJS: acute jaundice syndrome

A: Final selected model

B: ELISA sensitivity of 88% and specificity of 99% based on meta-analysis on the diagnostic accuracy of hepatitis E antibody tests (9).

C: RDT sensitivity of 93% and specificity of 99% based on lowest values in published literature (10,11),

D: PCR sensitivity and specificity set to same as ELISA

E: Combination of B, C, and D

**Supplemental Table 7. Characteristics of suspected hepatitis E cases with longitudinal (follow-up) test results.**

|  | | **Case type at enrollment** | | | | | | | |
| --- | --- | --- | --- | --- | --- | --- | --- | --- | --- |
| **Characteristic**  **N (col %)** | Overall N=363 | PCR+ RDT+ ELISA+  N=77 | PCR+ RDT+ ELISA-  N=0 | PCR+  RDT- ELISA+  N=5 | PCR+  RDT- ELISA-  N=1 | PCR- RDT+ ELISA+  N=15 | PCR-  RDT+  ELISA-  N=15 | PCR- RDT- ELISA+  N=7 | PCR-  RDT-  ELISA-  N=243 |
| Female | 172 (47·4) | 34 | - | 3 | 0 (0) | 5 | 8 | 3 | 119 |
| Age, median (IQR) | 15·5  (6·4, 23·7) | 9·9  (5·2, 16·0) | - | 6·6  (5·0, 10·5) | 10·6  (10·6, 10·6) | 14·3  (6·2, 16·9) | 24·8  (16·8, 31·6) | 6·6  (4·4, 15·5) | 18·5  (6·9, 25·4) |
| Age |  |  |  |  |  |  |  |  |  |
| 0-5 | 71 (19·6) | 17 (22·1) | - | 2 (40·0) | 0 (0) | 3 (20·0) | 0 (0) | 3 (42·9) | 46 (18·9) |
| 6-15 | 104 (28·7) | 37 (48·1) | - | 2 (40·0) | 1 (100) | 7 (46·7) | 2 (13·3) | 2 (28·6) | 53 (21·8) |
| 16-39 | 171 (47·1) | 20 (26·0) | - | 1 (20·0) | 0 (0) | 5 (33·3) | 11 (73·3) | 2 (28·6) | 132 (54·3) |
| 40+ | 17 (4·7) | 3 (3·9) | - | 0 (0) | 0 (0) | 0 (0) | 2 (13·3) | 0 (0) | 12 (4·9) |
| Days since jaundice onset, median (IQR) | 4·0  (3·0, 9·0) | 4·0  (3·0, 5·0) | - | 4·0  (3·0, 6·0) | 17·0  (17·0, 17·0) | 5·0  (4·5, 9·0) | 5·0  (3·5, 18·0) | 42·0  (9, 91) | 4·0  (3·0, 10·0) |
| Days since jaundice onset |  |  |  |  |  |  |  |  |  |
| ≤1 week | 256 (70·5) | 65 (84·4) | - | 4 (80·0) | 0 (0) | 10 (66·7) | 9 (60·0) | 2 (28·6) | 166 (68·3) |
| >1 week to 2 weeks | 48 (13·2) | 11 (14·3) | - | 0 (0) | 0 (0) | 3 (20·0) | 1 (6·7) | 1 (14·3) | 32 (13·2) |
| >2 weeks to 1 month | 30 (8·3) | 1 (1·3) | - | 1 (20·0) | 1 (100) | 1 (6·7) | 2 (13·3) | 0 (0) | 24 (9·9) |
| >1 month to 2 months | 13 (3·6) | 0 (0) | - | 0 (0) | 0 (0) | 0 (0) | 1 (6·7) | 2 (28·6) | 10 (4·1) |
| >2 months | 16 (4·4) | 0 (0) | - | 0 (0) | 0 (0) | 1 (6·7) | 2 (13·3) | 2 (28·6) | 11 (4·5) |
| Elevated ALT* | 59 (21·9) | 48 (73·8) | - | 1 (25·0) | 0 (0) | 1 (11·1) | 1 (9·1) | 0 (0) | 8 (4·6) |
| Elevated AST** | 22 (6·1) | 6 (30·0) | - | 0 (0) | ** | 1 (20·0) | 1 (12·5) | 0 (0) | 14 (14·0) |
| PCR positive at follow-up |  |  |  |  |  |  |  |  |  |
| Negative | 349 (96·1) | 64 (83·1) | - | 5 (100) | 0 (0) | 15 (100) | 15 (100) | 7 (100) | 243 (100) |
| Positive | 14 (3·9) | 13 (16·9) | - | 0 (0) | 1 (100) | 0 (0) | 0 (0) | 0 (0) | 0 (0) |
| IgM ELISA positive at follow-up |  |  |  |  |  |  |  |  |  |
| Negative | 277 (76·3) | 11 (14·3) | - | 2 (40·0) | 1 (100) | 3 (20·0) | 15 (100) | 4 (57·1) | 241 (99·2) |
| Positive | 82 (22·6) | 63 (81·8) | - | 3 (60·0) | 0 (0) | 12 (80·0) | 0 (0) | 3 (42·9) | 1 (0·4) |
| Indeterminate | 4 (1·1) | 3 (3·9) | - | 0 (0) | 0 (0) | 0 (0) | 0 (0) | 0 (0) | 1 (0·4) |
| Hospitalizations | 7 (1·9) | 1 (1·3) | - | 0 (0) | 0 (0) | 1 (6·7) | 0 (0) | 0 (0) | 5 (2·1) |
| Deaths | 1 (0·3) | 0 (0) | - | 0 (0) | 0 (0) | 0 (0) | 0 (0) | 0 (0) | 1 (0·4) |

*Missing for 94 suspected cases. For males, ALT ≥41 units per liter was considered elevated. For females, ALT >32 units per liter was considered elevated (23,24).

**Missing for 226 suspected cases. AST above 40 units per liter was considered elevated.

**Supplemental Table 8. Accelerated failure time models used to estimate median time from a positive test result at enrollment to a negative test result.** Grey shading indicates final selected model.

| **Test** | **Distribution** | **Median time to negative test**  **(95% CI)*** | **AIC** |
| --- | --- | --- | --- |
| PCR | Weibull  (Log-shape: 1·1, Log-scale: 3·1) | 20·3  (18·7, 22·5) | 91·7 |
|  | Log-normal  (Location: 2·9, Log-scale: -1·0) | 18·9  (16·9, 21·3) | 83·4 |
|  | Gamma  (Log-shape: 2·1, Log-scale: 0·94) | 19·3  (17·5, 22·0) | 85·5 |
| IgM ELISA | Weibull  (Log-shape: 0·56, Log-scale: 5·0) | 116·8  (83·8, 149·2) | 87·4 |
|  | Log-normal  (Location: 4·7, Log-scale: -0·06) | 114·6  (86·4, 182·5) | 88·2 |
|  | Gamma  (Log-shape: 0·77, Log-scale: 4·1) | 113·0  (87·4, 163·1) | 87·3 |

*Based on the 2·5^th^ and 97·5^th^ percentiles of 1,000 bootstrap draws

CI: confidence interval; AIC: Akaike Information Criterion
